# Fighting COVID-19 with Flexible Testing: Models and Insights

**DOI:** 10.1101/2020.11.17.20233577

**Authors:** Jiali Yu, Zuo-Jun (Max) Shen

## Abstract

Increasing testing capacities plays a substantial role in safely reopening the economy and avoiding a new wave of COVID-19. Pooled testing can expand testing capabilities by pooling multiple individual samples, but it also raises accuracy concerns. In this study, we propose a flexible testing strategy that adopts pooled testing or individual testing according to epidemic dynamics. We identify the prevalence threshold between individual and pooled testing by modeling the expected number of tests per confirmed case. Incorporating an epidemic model, we show pooled testing is more effective in containing epidemic outbreaks and can generate more reliable test results than individual testing because the reliability of test results is relevant to both testing methods and prevalence. Our study is the first to evaluate the interplay between pooled testing and a rapidly evolving outbreak to the best of our knowledge. Our results allay accuracy concerns about pooled testing and provide theoretical supports to empirical studies.

## Introduction

Testing plays a critical role in containing the COVID-19 outbreak and lifting containment measures, which have caused enormous economic losses and posed health risks, particularly among disadvantaged populations. However, increasing the testing capability to meet the demand and improving the reliability of test results remains to be challenging tasks.

Pooled testing is to test samples drawn from multiple individuals as a pooled sample, also called a pool^1^. If a pool tests negative, everyone in this pool is considered negative. If the result is positive, which indicates that at least one individual in this pool is infected, then all the samples need to be retested individually. Prevalence, test accuracy, and pool size all have great impacts on pooled testing. When a large proportion of pools test negative, pooled testing can clear groups of patients using fewer assays. Conversely, if most pools yield positive results, pooled testing may sacrifice accuracy to improve efficiency.

Although pooled testing provides an alternative to expand the testing capability, policymakers have remained conservative about pooled testing, such as mandating a small pool size when the positive result rate is low. For example, the Indian Council of Medical Research suggested pooling five specimens for COVID-19 testing in communities where the positive rate was less than 5%^2^. The U.S. Food and Drug Administration (FDA) has approved only one COVID-19 diagnostic test for pooling up to four individual samples^3^. However, recent empirical studies have shown that the pool size could be expanded to 32 nasopharyngeal swabs while maintaining acceptable test accuracy^4^, and pools of 20 saliva samples help control the epidemic when the prevalence is less than 1%^5^.

Recent studies mainly focused on investigating efficiency gains and the accuracy losses of different pooled algorithms^4-17^, such as multi-stages and multi-dimensions procedures. In this study, we focus on Dorfman’s classic two-stage procedure because it is the most utilized algorithm in practice. Previous research mainly focuses on improving efficiency, i.e., minimizing the expected number of tests per subject or the total number of tests. Aprahamian, Bish, and Bish are the first to incorporate testing accuracy into pooled testing^18,19^. Moreover, Aprahamian, Bish, and Bish analytically characterize key structural properties and derive the threshold policy of optimal Dorfman’s pool size for the first time^19^. Their results show that pooled testing is always better than individual testing when the test sensitivity is imperfect. However, when the prevalence is higher than the threshold, the efficiency gains of pooled testing mainly come from the false negative results in first the round of testing, which avoid the second round of testing. During a pandemic, flexible testing should choose individual or pooled testing by trading off efficiency and accuracy. Moreover, during a pandemic, the interplay between testing and fast-changing prevalence remains unclear. The insufficient testing capacity and inaccurate testing technologies weaken the mitigation effects of testing and the reliability of test results.

We aim to provide a broad picture of testing in the pandemic from a public health policy perspective. We propose a flexible testing strategy that adopts pooled testing and individual testing according to epidemic dynamics. Based on previous research of optimal pooled testing^19^, we model the expected number of tests per confirmed cases and identify the prevalence threshold to determine when testing strategies must be changed. We use an epidemic model to compare flexible and individual testing in population screening. Our results contradict the common belief that pooled testing is less accurate than individual testing and that only highly accurate tests facilitate the safe reopening of the economy. We show that flexible testing is more cost-effective for containing the outbreaks and can generate more reliable results than individual testing. The higher capacity and frequency achieved by appropriately adopting a pooled testing strategy or newly developed tests ease the containment measures and prevent infections, even if tests have a low ability to detect the virus.

## Results

### Flexible testing strategy

The objective of testing during a pandemic is to identify and remove infected patients from the transmission chain as soon as possible. Based on the performance measures derived by Aprahamian, Bish, and Bish^18,19^, we measure the overall efficiency and effectiveness of testing by the expected number of tests required to identify an infected patient, defined as the cost-performance ratio (CPR). Pool size *n* = 1 represents individual testing. *n* > 1 represents pooled testing. CPR depends on the prevalence *p*, pool size *n*, test sensitivity *S*_*e*_ (true positive rate), and specificity *S*_*p*_ (true negative rate) as follows (see methods for details on the derivation of the CPR):

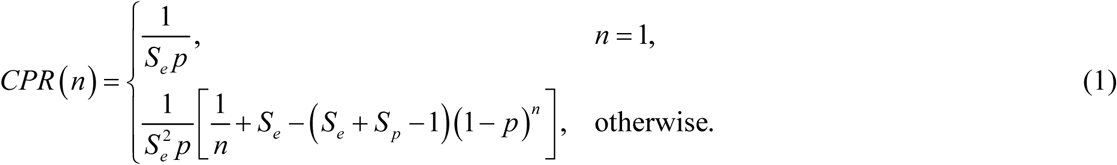

We derive the flexible testing strategy that adopt individual or pooled testing according to epidemic dynamics, based on the threshold policy of the optimal pooled testing^19^(see more details in Methods section and Supplementary Material). During the pandemic, the flexible testing strategy with an optimal pool size *n*^*^ that minimizes the CPR follows:

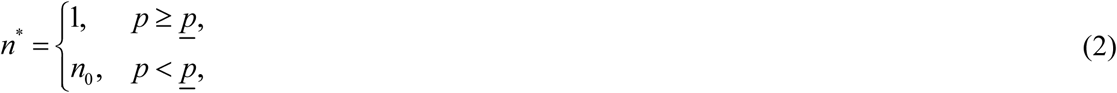

where 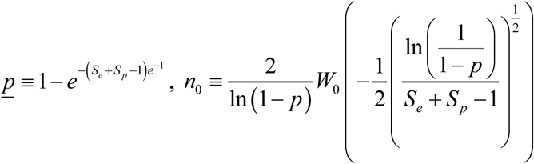 have been derived in previous research on optimal pooled testing in terms of the expected number of tests per subject^19^. *W*_0_ (.) is the principal branch of Lambert W function^20^.

Compared with previous research on optimal pooled testing^19^, our research focuses on comparing individual and pooled testing, in terms of *CPR*(*n*). If the prevalence is lower than the threshold, pooled testing with pool size *n*_0_ is preferred^19^. Otherwise, individual testing (*n*^*^ = 1) is the optimal strategy.

Moreover, *n*^*^, *CPR* (*n*^*^), and *<u>p</u>* have the simple monotonicity that offers clear directions for decision-makers to modify testing strategies to identify an infected patient with minimum tests. The results are as follows (see proof in supplementary material):

1. The prevalence threshold increases in sensitivity and specificity, as shown in Figure 1A. If the testing sensitivity and specificity are 70% and 95%, the prevalence threshold between individual and pooled testing is 21.2%. If tests were perfect, the prevalence threshold is up to 30%, and we can apply pooled testing in areas with higher prevalence.
2. The optimal pool size monotonically decreases with the increasing prevalence, sensitivity, and specificity^19^, as shown in Figure 1B, D. The optimal pool size varies more with prevalence than with test sensitivity and specificity, especially when prevalence is lower than 1%. When the prevalence is higher than 1%, the optimal pool size changes no more than one sample for a 10% fluctuation of sensitivity or specificity.
3. The CPR of optimal pool size decreases with increasing prevalence, sensitivity, and specificity, as shown in Figure 1C, E. As more people get infected, or we have more accurate testing technology, we can identify infected patients with fewer tests.

**Figure 1.**
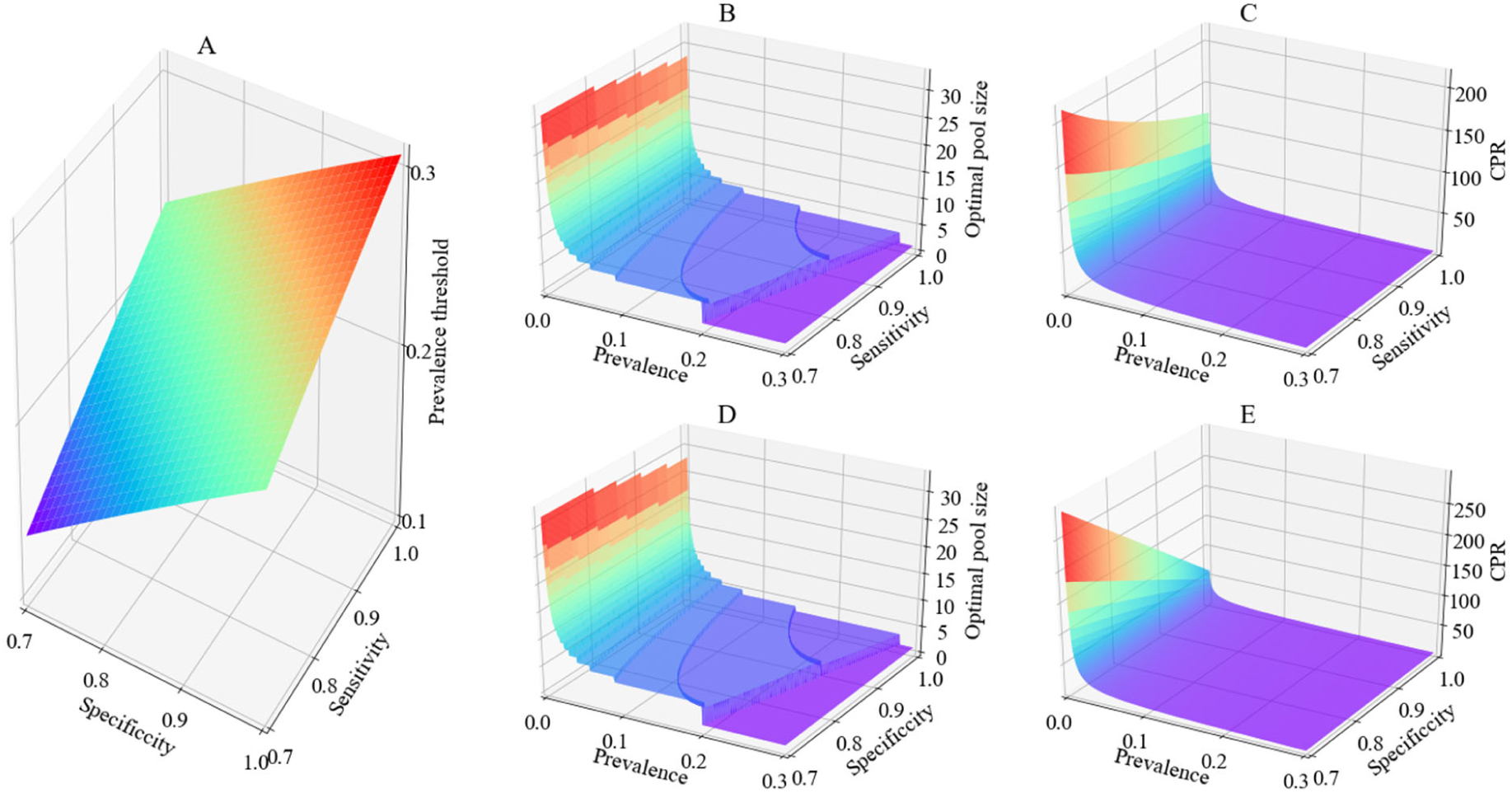
Monotonicity of the prevalence threshold, optimal pool size, and CPR. (A): The prevalence threshold is monotonically increasing in sensitivity and specificity. (B) and (C): the optimal pool size and the corresponding CPR is monotonically decreasing in prevalence and sensitivity, assuming the specificity is 90%. (D) and (E): the optimal pool size and the corresponding CPR is monotonically decreasing in prevalence and specificity, assuming the sensitivity is 90%. The prevalence ranges from 0.15%–30%.

### Application of flexible testing strategy

We illustrate the application of flexible testing using testing data from the United States. Figure 2 shows the seven-day average positive rate, the optimal pool sizes, and the CPRs of different testing strategies for 50 U.S. states and Washington, D.C. Based on testing data from the COVID Tracking Project https://covidtracking.com/, between October 28–November 2, 2020, five states reported a seven-day average positive rate higher than 20%, namely, South Dakota (50.6%), Iowa (37.4%), Kansas(36.0%), Idaho (33.4%), and Wyoming(30.4%) (figure 2A). Note that positive rates are higher than prevalence rates where testing capacity is limited because hospitals prioritize testing individuals with symptoms or a higher risk of infection. For simplicity, we use test positive rates to represent the prevalance in the tested population.

**Figure 2:**
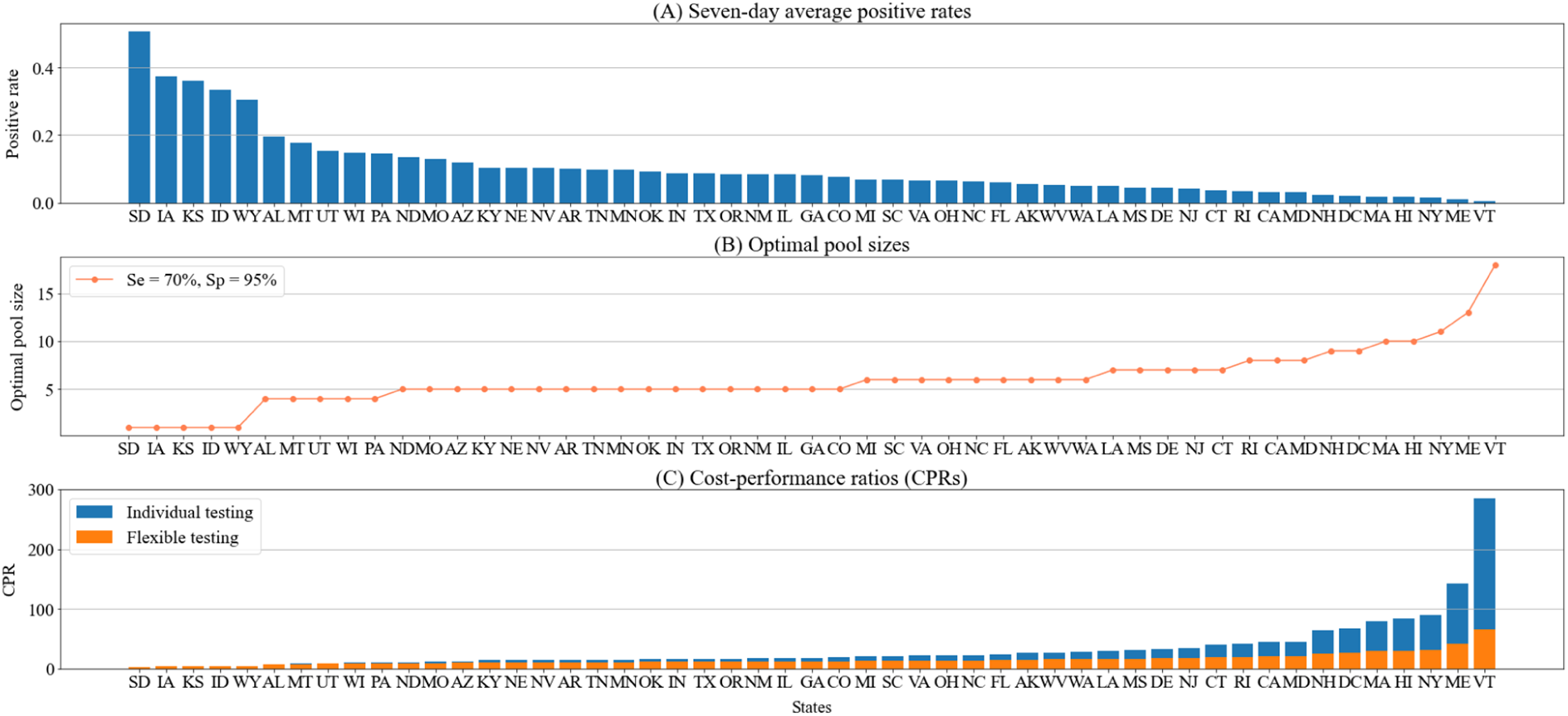
COVID-19 testing in the United States. (A) The seven-day average positive rate of 50 U.S. states and Washington, D.C., between October 28–November 2, 2020. (B) The optimal pool size of the flexible testing strategy. (C) The CPRs of the flexible and individual testing strategy. We assume that sensitivity and specificity are 70% and 95%, respectively.

If the test sensitivity is 70% and specificity is 95%, which are reasonable estimates of main diagnostic test accuracy (involving a nasopharyngeal swab)^21^, 45 states and Washington, D.C., where positive rates are lower than 21.2%, can apply pooled testing to increase testing capabilities. The maximum pool size approved by the U.S. FDA is four individual samples, which is the optimal pool size for five states where positive rates are 12.5%–21.2%. 40 states and Washington, D.C., with lower than 12.5% positive rates, can implement larger pool sizes than four to increase testing capabilities further. The current limitation on the maximum pool size is too conservative and restraints the effectiveness of pooled testing. When the prevalence is less than 1%, flexible testing saves more than 70% tests to identify an infected patient than individual testing. Taking Vermont as an example, the positive rate is 0.5%. Flexible testing with the pool size of 18 takes 66 tests to identify an infected patient, while individual testing takes 286 tests. Flexible testing can alleviate the current shortage of coronavirus tests.

### Comparison of individual and flexible testing strategies

A flexible testing strategy should consider the prevalence rate, which changes over time among different groups. In practice, we can implement targeted testing tactics by categorizing people into different risk groups based on their local prevalence rates, symptoms, and exposure histories. In this study, we focus on the change of the prevalence rate over time. We use a susceptible–infected–removed (SIR) model to compare individual and flexible testing strategies in population screening considering different containment measures, test sensitivities, specificities, and capacities.

In baseline scenarios, the initial prevalence is 1%. The testing sensitivity and specificity are 73.3% and 98.6%, respectively. Limited testing capacity allows individually test all population within 100 days. Containment measures suppress the reproductive number to 1.5. As shown in Figures 3A and 3B, the total infection rate increases exponentially without testing and more drastic containment measures. Individual testing only flattens the curve. Flexible testing contains the outbreak more effectively and causes fewer infections than individual testing.

**Figure 3:**
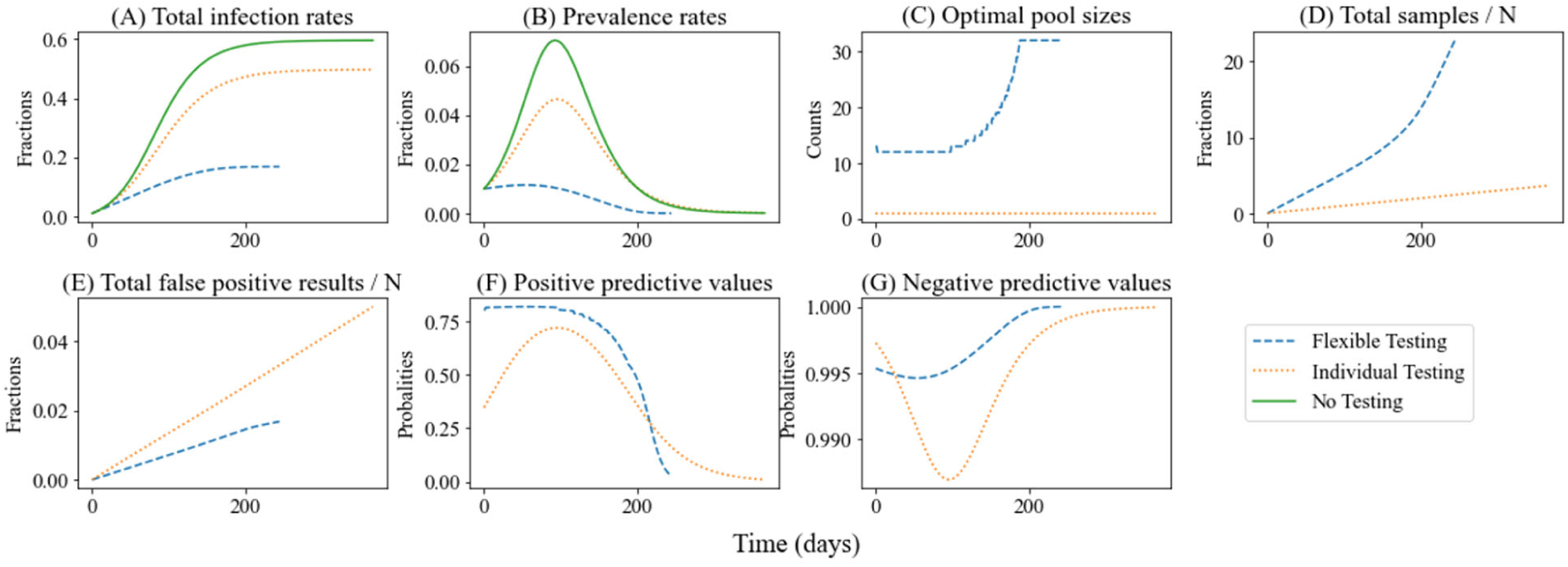
Epidemic dynamics and test performances of individual and flexible testing.

Because the prevalence is lower than the threshold, flexible testing adopts pooled testing to increase testing capabilities. As the prevalence in pooled testing decreases (Figure 2B), the optimal pool size increases until it reaches the upper limit (Figure 2C). Pooled testing increases testing capabilities make it possible to test people more frequently. In baseline scenarios, pooled testing tests all population five times within the initial 100 days, while individual testing only tests all population once. Although pooled testing provides more results than individual testing (Figure 2D), it generates fewer false positive results (Figure 2E), avoiding unnecessary treatments and isolations.

We measure the reliability of the test results by positive predictive values (PPVs) and negative predictive values (NPVs). PPVs and NPVs incorporate information about the prevalence, pool size, test sensitivity, and specificity (see the supplementary material for details of the PPV, NPV, sensitivity, and specificity). High PPVs and NPVs correspond to reliable positive and negative results, respectively.

Contrary to the common belief that pooled testing is less accurate than individual testing, we show that pooled testing can generate more reliable results because the reliability of test results is relevant to testing methods and prevalence. More precisely, pooled testing can be associated with higher PPVs during a pandemic (Figure 2F) because each positive sample is assayed twice, include a pooled test and an individual retest. On the other hand, the NPVs of pooled testing are initially lower than those of individual testing, but they quickly increase and become higher than those of individual testing (Figure 2G) when the prevalence rate under pooled testing is significantly lower than that under individual testing (Figure 2B). However, research that assumes a fixed prevalence shows that pooled testing always leads to lower NPVs than those of individual testing ^22^.

During the initial stage of the outbreak, we faced great challenges in designing, developing, and expanding the diagnostic tests. Drastic containment measures, such as a complete lockdown, effectively suppressed the total infection rate but halted most economic activities. Flexible testing offers an alternative to ramp up testing capabilities within a short time and accelerate the suppression of outbreaks (Figure 4A, B). With the lifting of containment measures (Figure 4C, D), the effect of flexible testing approaches that of individual testing, only mitigating the outbreak. If we let the virus spread with few containment measures, the total infection rate will increase exponentially and surpass 90%, and limited tests have little effect on containing the outbreak (Figure 4D). Containment measures and testing should be coordinated to reduce both infections and economic losses. To keep total infection rates lower than the same level, flexible testing requires less stringent containment measures than individual testing.

**Figure 4:**
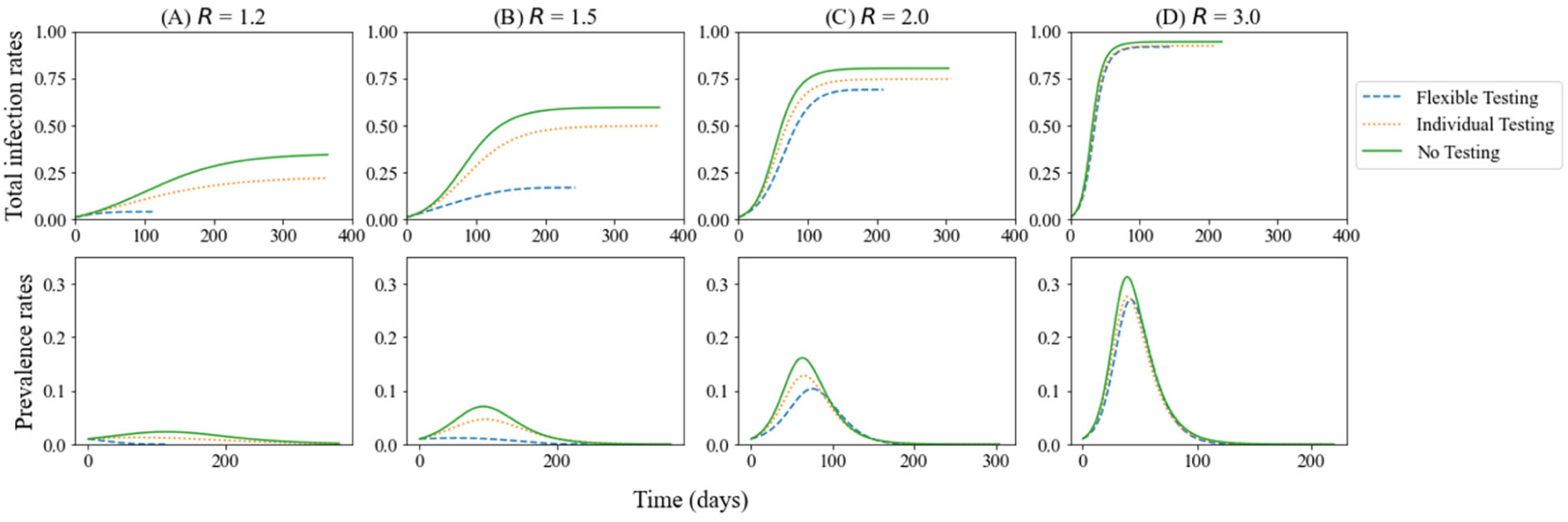
Effects of testing under different containment measures. Smaller *R* implies more stringent containment measures. The top and bottom show total infection rates and prevalence rates, respectively.

Test sensitivity and specificity affect epidemic dynamics differently under different testing strategies, as shown in Figure 5. Test sensitivity impacts epidemic dynamics and test performances for both individual and pooled testing. Higher test sensitivity reduces total infection rates (Figure 5A) and increases NPVs (Figure 5B), which imply more reliable negative results.

**Figure 5:**
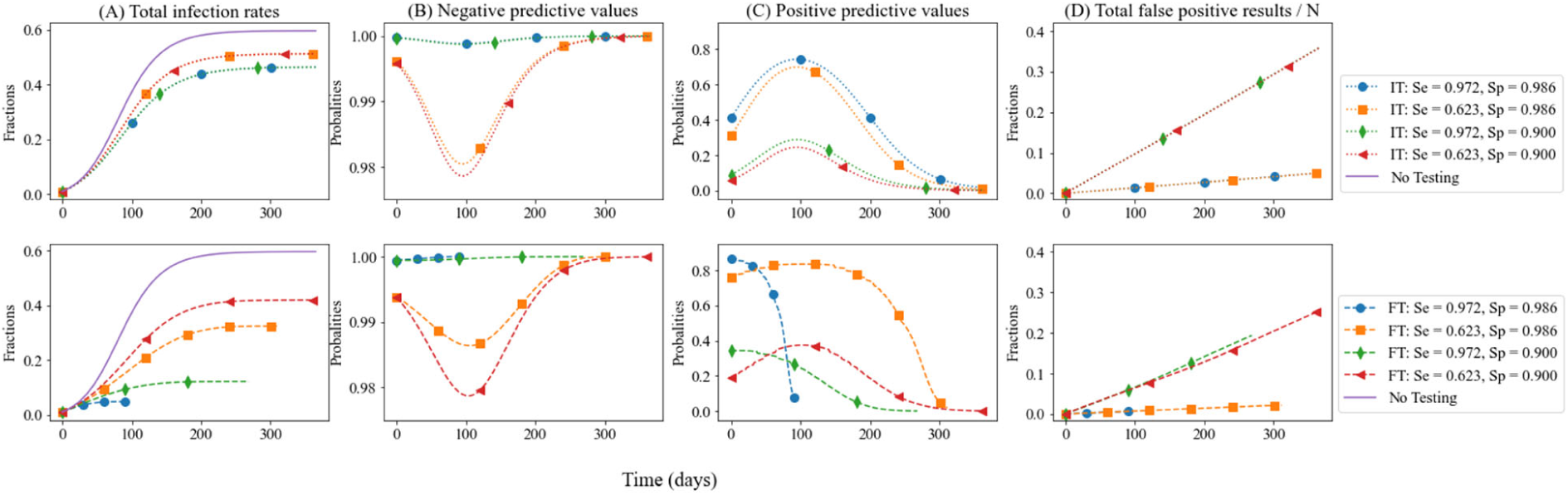
Comparison of different testing strategies and technologies. The top and bottom rows show individual testing (IT) and flexible testing (FT), respectively. Testing technologies vary in sensitivity *S*_*e*_ and specificity *S*_*p*_, as indicated.

Specificity has a negligible influence on epidemic dynamics under individual testing (Figure 5A top). However, we show that specificity is also important for pooled testing. Lower specificity causes more false positive results of pooled samples in the first stage and then leads to more unnecessary individual retests. Increasing testing sensitivity or specificity can dramatically strengthen the effectiveness of pooled testing on containing outbreaks (Figure 5A bottom). Moreover, higher test specificity reduces false positive results (Figure 5D) and increases PPVs (Figure 5C), which imply more reliable positive results.

The testing capacities achieved with different technologies vary significantly. In-home rapid tests enable the possibility of screening a population frequently. When the initial prevalence is 5% and there are few containment measures (R=3), individual testing twice a week can reverse the outbreak and keep the total infection rate no more than 20%, even using an inaccurate testing technology (sensitivity: 62.3% and specificity: 90%) (Figure 6A, B). This result contradicts the common belief that highly accurate tests are prerequisites to safely reopening the economy. As the testing frequency increases, inaccurate technologies reduce infections (Figure 6A, B); however, FPs rapidly increase because of the low specificity (Figure 6C).

**Figure 6:**
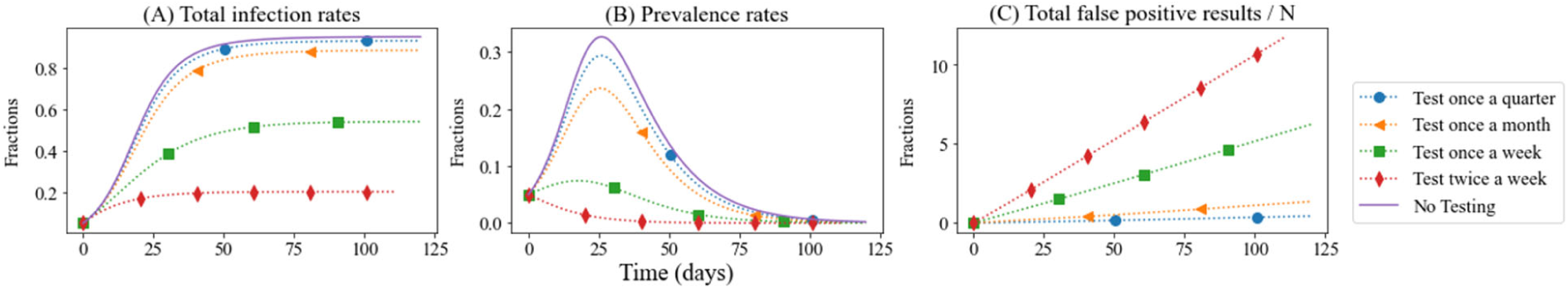
Comparison of different testing frequencies of individual testing. These scenarios show total infection rates (A), prevalence rates (B), and false positives (C) of individual testing with an initial prevalence of 5% using an inaccurate technology (62.3% sensitivity and 90% specificity). The reproductive number is 3. We compare the quarterly, monthly, weekly, and twice-weekly testing frequency.

Frequent testing, such as the twice-weekly testing, could enable the economy to reopen while maintaining the total infection rate much lower than the herd immunity level. Compared with the devastating economic cost of containment measures, increasing testing capacity is more cost-effective in containing the outbreak. Our results are consistent with the findings of the previous study that increasing testing frequency contributes to a safer reopening, even if tests are inaccurate^23^.

## Discussion and conclusion

Flexible testing is more cost-effective in containing the outbreaks and can generate more reliable results than individual testing during the pandemic. We identify the prevalence threshold between individual and pooled testing in terms of CPR. Simulation results show that increasing testing capability is a more feasible strategy to expedite identifying infected patients than improving testing sensitivity and specificity in short term. Moreover, pooled testing can increase the reliability of negative results by lowering the prevalence and improve the reliability of positive results by testing positive samples twice.

Our analysis also subjects to several limitations. First, we assume the prevalence is known, and then choose individual or pooled testing according to the prevalence. During the pandemic, developing methods to estimate the prevalence of COVID-19 is crucial for adjusting containment policies. Moreover, we use a well-mixed SIR model to capture the difference between individual and flexible testing. More detailed transmission models that consider the heterogeneous population and social contact patterns can provide a more specific testing performance evaluation. In addition, assessing the diagnostic accuracy of tests administered to pooled samples and asymptomatic individuals is also an urgent priority^5,24^.

Flexible testing and the newly developed tests, which are low cost and noninvasive, make widespread and frequent testing accessible and help navigate uncertainty after the reopening. Compared with the painful containment measures, increasing testing frequency enables people to return to workplaces and students to go back to schools more safely. For example, the U.S. National Football League is individually testing all players and team personnel every day or every other day during the regular season except game day because false positives may cause the delay or cancellation of a game. The seven-day positive rate is only 0.017% between August 30– September 5, 2020^25^. Pooled testing can save more than 95% of tests for the National Football League. Moreover, the University of Illinois Urbana-Champaign (UIUC) is testing students twice-weekly using rapid saliva tests^26^. Between July 6–November 14, 2020, the daily positive rate is no more than 2.86% (August 30, 2020). As these positive rates are much lower than the threshold, compared with individual testing, pooled testing can save substantial tests and reduce numerous false positives by testing each positive sample twice. For example, the University of Cambridge and the University of Nottingham are implementing pooled testing to keep campuses open. Meanwhile, healthcare providers should offer more information on interpreting test results and organize timely follow-up testing to rule out false positive and false negative results.

Countries are striving to increase their testing capabilities to control the COVID-19 outbreak and safely reopen their economies. We suggest governments and agencies increase the testing capability and frequency by the approval of larger pool sizes and more fast test technologies. As we learn more about the pandemic by testing more people and conducting more research, we will gain confidence and win the fight against COVID-19.

## Methods

### Flexible testing strategy

Containing the epidemic outbreak requires both high testing efficiency and accuracy. The faster the infected patients are removed from the transmission chain, the fewer people will become infected later. Although test sensitivity and specificity measure the intrinsic accuracy of a test in identifying patients with and without the disease, prevalence-dependent test performance measures are more practical in comparing testing strategies under different disease prevalences *p*∈(0,1). Because the test is imperfect and healthy people have a higher probability of receiving negative test results than infected people, we assume *S*_*e*_ + *S* _*p*_ − 1∈ (0,1).

We first review the performance measures derived by Aprahamian, Bish, and Bish, including the per-subject expected number of tests 𝔼[*T* (*n*)] and the per-subject expected number of false negatives 𝔼[*FN* (*n*)] ^18,19^; that is,

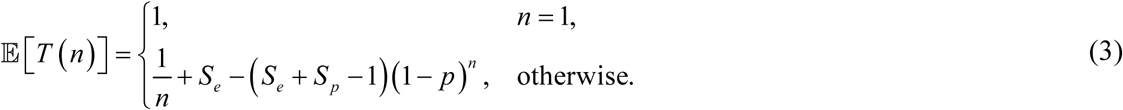

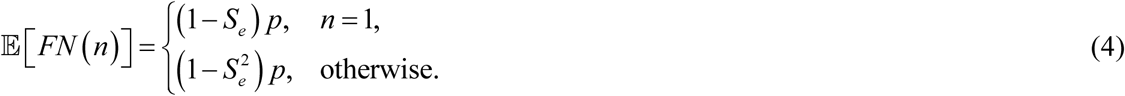

The per-subject expected number of true positives 𝔼[*TP* (*n*)] is given by Equation (5), because 𝔼[*TP* (*n*)] + 𝔼[*FN* (*n*)] = *p*.

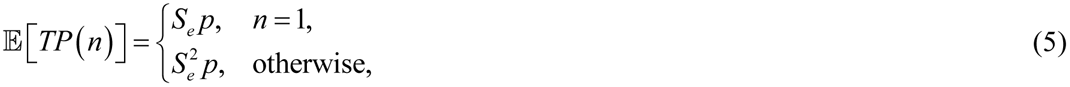

We extended previous research on optimal pooled testing^19^ by modeling the cost-performance ratio (CPR), the ratio of the per-subject expected number of tests to the per-subject expected number of true positives. CPR represents the number of tests per confirmed case, a commonly used statistic evaluating the overall performance of testing in the COVID-19 pandemic. CPR incorporates 𝔼[*TP* (*n*)] to capture the accuracy concerns over false negative results, which might give people a false sense of security to break the social distancing orders and lead to widespread transmission.

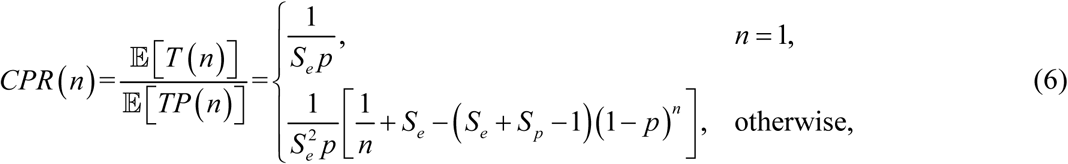

Note that minimizing *CPR*(*n*) is the same as minimizing the expected number of tests per subject 𝔼[*T* (*n*)] for pooled testing^19^. Aprahamian, Bish, and Bish have proved the optimal pooled testing in terms of 𝔼[*T* (*n*)]. Thus, we can prove the flexible testing strategy by comparing individual testing with optimal pooled testing (see supplementary material for more details). We further discuss the flexible testing strategy under the realistic scenario in the supplementary material.

### Epidemic Model

The SIR model captures the prevalence variation over time. Susceptible individuals *S*_*t*_ are infected through contact with unidentified infected individuals *I*_*t*_. The removed people *R*_*t*_ consist of recovered people and confirmed cases who are isolated until they pose no risk. Although it seems more natural to utilize population counts, we use fractions of the total population *N* to simplify the discussion and calculation, so that *S*_*t*_ + *I*_*t*_ + *R*_*t*_ =1. Therefore, the prevalence is *I*_*t*_. The epidemic dynamics are described by the following system of ordinary differential equations:

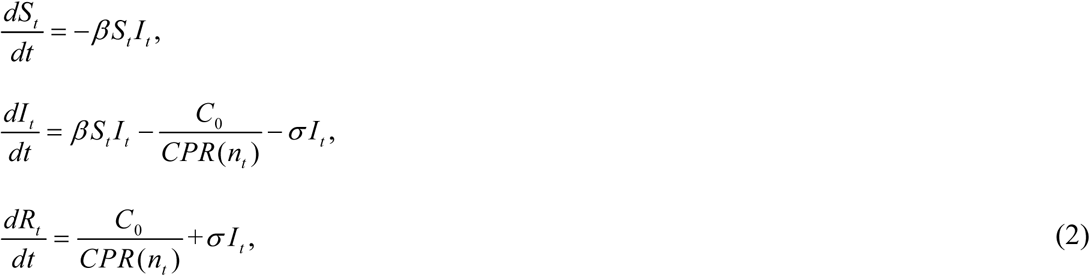

where *β* denotes the transmission rate, and *σ* is the recovery rate. To prevent infections as many as possible, we assume that decision-makers make full use of daily testing capacity *C*_0_. The number of infected individuals who are isolated after testing positive is *C*_0_ / *CPR*(*n*_*t*_), where *n* _*t*_ is the pool size during time interval *dt*. For flexible testing strategy, the optimal pool size 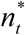 is chosen dynamically according to the prevalence *I*_*t*_ to minimize the *CPR*(*n*_*t*_). Individual testing test one sample per assay all the time (i.e., *n*_*t*_ =1).

### Simulation Settings

We compare the epidemic dynamics and performances of individual and flexible testing strategies in a pandemic by simulating different containment measures, testing sensitivity, specificity, and capacity. The baseline values and the value ranges used for sensitivity analyses are listed in Table 1. We end the population screening when the prevalence is lower than 10^_5^.

**Table 1.** Model parameter values at baseline and the ranges used for sensitivity analyses.

| Parameter | Baseline Value | Range | Details and references |
| --- | --- | --- | --- |
| Maximum pool size $M$ | 32 | | A recent study revealed that the COVID-19 real-time quantitative polymerase chain reaction (RT-qPCR) test sensitivity retained 90% for a batch of up to 32 nasopharyngeal swabs<sup>4</sup>. |
| Recovery time $\sigma^{-1}$ | 14 | | The median recovery time for mild cases is approximately two weeks<sup>27</sup>. |
| Reproductive number $R$ with containment measures | 1.5 | 1.2, 1.5, 2.0, 3.0. | The reproductive number can be thought of the expected number of new infections that are generated directly from one infected individual. We calculate $\beta$ as $\beta = R \cdot \sigma$. We use $R$ to capture the overall impact of containment measures. A smaller $R$ corresponds to more stringent measures, which cause more economic losses. |
| Sensitivity $S_e$ | 0.733 | 0.623, 0.972. | The test accuracy varies because of human error and variability in specimens. We use the nasopharyngeal swab PCR test as the |
| Specificity $S_p$ | 0.986 | 0.900,<br>0.986. | baseline. We analyze the main range of sensitivity and specificity of PCR tests (involving the nasopharyngeal swab, saliva, and sputum samples) based on a systematic review of COVID-19 test accuracy<sup>28</sup>. |
| Testing capacity $C_0 / N$ | 0.010 | 0.011,<br>0.033,<br>0.143,<br>0.286. | Testing capacity $C_0 = 0.01N$ represents that we can individually test $N$ people within 100 days. Sensitivity analysis compare the quarterly, monthly, weekly, and twice-weekly testing frequency under individual testing. |
| Initial prevalence $I_0$ | 1% | 5% | The initial prevalence is 1% of people are infected with COVID-19 but not removed from the transmission chain. |

## Data Availability

We use the testing data provided by the COVID Tracking Project who collects, cross-checks, and publishes COVID-19 testing data of the United States. To view more descriptions about data, please visit https://covidtracking.com/about-data. Johns Hopkins Coronavirus Resource Center provides a visualization tool for testing outcomes https://coronavirus.jhu.edu/testing/individual-states. The on-campus COVID-19 testing data dashboard of UIUC is available at https://go.illinois.edu/COVIDTestingData. All code is available for free use at https://github.com/JL-YU/Flexible-Testing.

https://coronavirus.jhu.edu/testing/individual-states

https://go.illinois.edu/COVIDTestingData

https://github.com/JL-YU/Flexible-Testing

## Contributors

Jiali Yu developed the model and prepared the first draft of the paper. Zuo-Jun (Max) Shen supervised the study, contributed to data interpretation. All authors contributed to writing the manuscript, verified the underlying data, and approved the final version..

## Supplementary Information

## Supplementary Methods

### Proof of Flexible Testing Strategy

Flexible testing strategy points out the prevalence threshold of individual and pooled testing. Figure 1 shows the *CPR*(*n*) curve and flexible testing strategy under different prevalence ranges.

**Figure 1:**
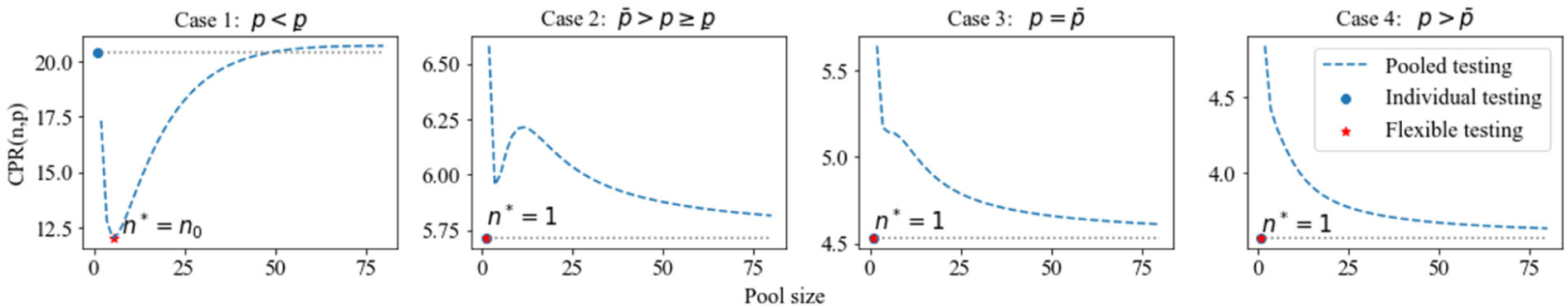
CPR curve and flexible testing strategy under different prevalence ranges. The red star shows the flexible testing strategy and corresponding CPR. To summarize all cases, we have: if *p* < *<u>p</u>*, the global minimizer of *CPR*(*n*) is *n*^*^ = *n*_0_ ; if *p* ≥ *<u>p</u>*, the global minimizer of *CPR*(*n*) is *n*^*^ = 1, i.e., flexible testing should choose individual testing.

The proof of flexible testing strategy is based on the optimal pooled testing. We refer interested readers to reference 1. Aprahamian, Bish, and Bish are the first to derive the closed form solution of optimal pooled size 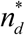, as shown in Equation (1), by minimizing per-subject expected number of tests 𝔼[*T* (*n*)] ^1^.

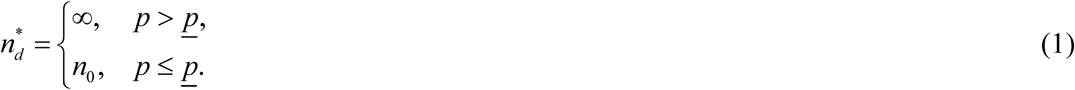

The optimal pooled testing, in terms of 𝔼[*T* (*n*)], follows a threshold policy: if the prevalence is higher than the threshold *<u>p</u>*, the optimal pool size should be as large as possible; otherwise, the optimal pool size is *n*_0_ ^1^.

If we only consider pooled testing, *CPR*(*n*) and 𝔼[*T* (*n*)] have the same optimal pool size because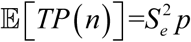 is independent with pool size *n*. Thus, we can prove the flexible testing strategy by comparing individual testing with optimal pooled testing in terms of *CPR*(*n*).

Step 1: Compare 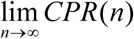 with 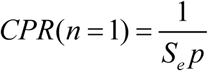.

Reference 1 has approved 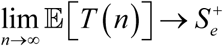 which represents 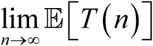 approaches *S* _*e*_ from above^1^. Thus, we have:

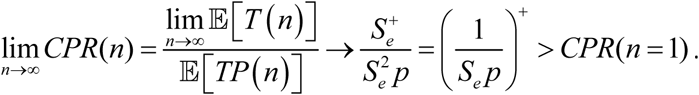

As 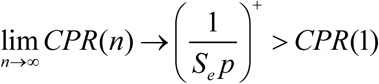, individual testing can always have a lower CPR than pooled testing with an infinitely large pool size.

Step 2: Compare *CPR*(*n*_0_) with 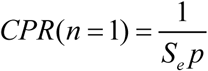.

When *p* ≤ *<u>p</u>*, Aprahamian, Bish, and Bish have already proved that 𝔼[*T* (*n*_0_)] ≤ *S*_*e*_ and *n*_0_ ≥ *e* ^1^. Based on their results, when *p* < *<u>p</u>*, we have that:

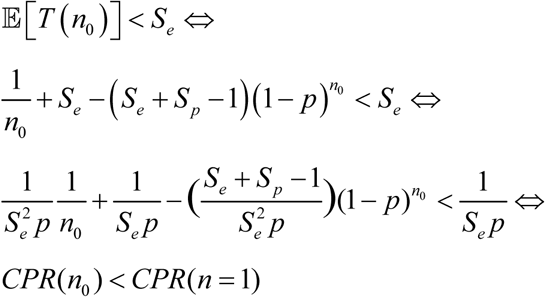

Thus, when *p* < *<u>p</u>*, pooled testing with pool size *n*_0_ has lower CPR than individual testing. When *p* = *<u>p</u>*, we have *CPR*(*n*_0_) = *CPR*(*n* = 1). In this scenario, we suggest individual testing which is simpler than pooled testing. Moreover, the integer pool size in practice leads to a smaller CPR. We will discuss the flexible testing strategy considering realistic scenarios at the end of the supplementary material.

To summarize the threshold policy of individual and pooled testing in terms of CPR: if *p* < *<u>p</u>*, pooled testing with optimal pool size *n*_0_ is better than individual testing; if *p* ≥ *<u>p</u>*, individual testing outperforms pooled testing.

### Monotonicity

In this part, we derive and prove the monotonicity properties of prevalence threshold, optimal pool size, and CPR.

#### Property 1

The prevalence threshold *p* between individual and pooled testing is increasing in *S*_*e*_ and *S* _*p*_.

#### Proof of Property 1

The first order derivatives of prevalence threshold in *S* _*e*_ and *S* _*p*_ are:

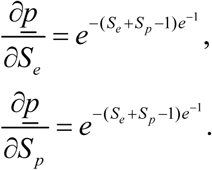

Because 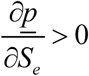 and 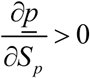, *<u>p</u>* is increasing in *S*_*e*_ and *S* _*p*_.

#### Property 2

*n*^*^ is non-increasing in *p, S* _*e*_, and *S* _*p*_.

Property 2 has been proved by Aprahamian, Bish, and Bish, when *p* < *<u>p</u>* ^1^. When *p* ≥ *<u>p</u>*, flexible testing adopts individual testing, i.e., *n*^*^ = 1. As *n*_0_ ≥ *e* > 1^1^, Property 3 holds for all prevalence rate.

#### Property 3

*CPR*(*n*^*^) is decreasing in *p, S*_*e*_, and *S* _*p*_.

#### Proof of Property 3

1. Defining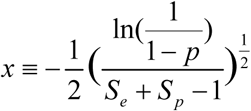 the derivative of *CPR*(*n\**) with respect to *p* is given by:

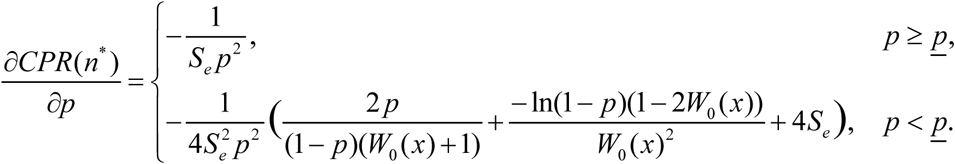 When 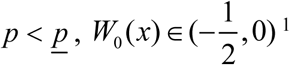. We have 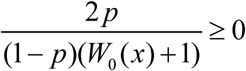 and 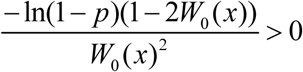. Then, we have 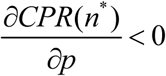 Therefore, *CPR*(*n*^*^) is decreasing in *p*.
2. The derivative of *CPR*(*n*^*^) with respect to *S* _*e*_ is given by:

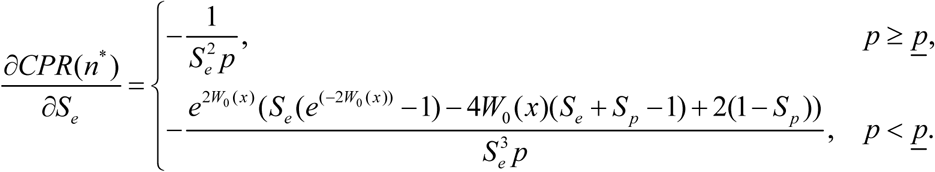 When 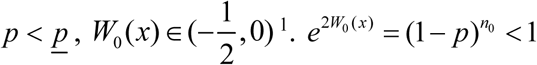, and (*S*_*e*_ + *S*_*p*_ − 1). We have 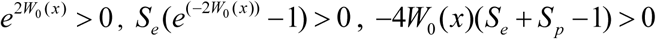, and 2(1 − *S* _*p*_) > 0. Thus, we have 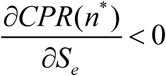. Therefore, *CPR*(*n\**) is decreasing in *S*_*e*_.
3. The derivative of *CPR*(*n*^*^) with respect to *S* _*p*_ is given by:

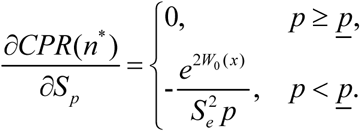 Because 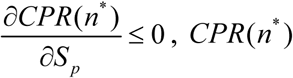 is non-increasing in *S* _*p*_.

### Flexible testing under realistic scenarios

Using the same assumptions for realistic scenarios as in the previous research of optimal pooled testing, the pooled testing has an upper limit on pool size *M* ∈ 𝕫_+_ to confine dilution effect^1^. We can ignore dilution effect if pool size is lower than *M*. Moreover, pool size should be an integer in practice. Corollary 1 shows the flexible testing under realistic scenarios.

#### Corollary 1

The flexible testing strategy under realistic scenario follows:

1. If *p* ≥ *p, n*^*^ = 1.
2. If 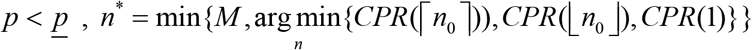.

According to Corollary 1, we can find the optimal integer pool size with negligible dilution effect. It should be stressed here that all monotonicity we discussed above holds for the realistic scenario. We use the realistic optimal pool size to derive all the results and conclusions.

## References

1. Dorfman R. The Detection of Defective Members of Large Populations. The Annals of Mathematical Statistics 1943; 14(4): 436–40. http://www.jstor.org/stable/2235930.

2. Indian Council of Medical Research. Advisory on feasibility of using pooled samples for molecular testing of COVID-19, 2020. https://www.icmr.gov.in/pdf/covid/strategy/Advisory_on_feasibility_of_sample_pooling.pdf.

3. FDA. Coronavirus (COVID-19) Update: FDA Issues First Emergency Authorization for Sample Pooling in Diagnostic Testing, 2020. https://www.fda.gov/news-events/press-announcements/coronavirus-covid-19-update-fda-issues-first-emergency-authorization-sample-pooling-diagnostic.

4. Yelin I, Aharony N, Tamar ES, et al. Evaluation of COVID-19 RT-qPCR Test in Multi sample Pools. Clinical Infectious Diseases 2020. DOI: 10.1093/cid/ciaa531.

5. Watkins AE, Fenichel EP, Weinberger DM, et al. Pooling saliva to increase SARS-CoV-2 testing capacity. medRxiv 2020: 2020.09.02.20183830. DOI: 10.1101/2020.09.02.20183830.

6. Augenblick N, Kolstad JT, Obermeyer Z, Wang A. Group Testing in a Pandemic: The Role of Frequent Testing, Correlated Risk, and Machine Learning. National Bureau of Economic Research Working Paper Series 2020; No. 27457. DOI: 10.3386/w27457.

7. Majid F, Omer SB, Khwaja AI. Optimising SARS-CoV-2 pooled testing for low-resource settings. The Lancet Microbe 2020; 1(3): e101–e2. DOI: 10.1016/S2666-5247(20)30056-2.

8. Libin PJK, Willem L, Verstraeten T, Torneri A, Vanderlocht J, Hens N. Assessing the feasibility and effectiveness of household-pooled universal testing to control COVID-19 epidemics. medRxiv 2020: 2020.10.03.20205765. DOI: 10.1101/2020.10.03.20205765.

9. Deckert A, Bärnighausen T, Kyei N. Pooled-sample analysis strategies for COVID-19 mass testing: a simulation study. Bulletin of the World Health Organization 2020; 98: 590–8. DOI: 10.2471/BLT.20.257188.

10. Praharaj I, Jain A, Singh M, et al. Pooled testing for COVID-19 diagnosis by real-time RT-PCR: A multi-site comparative evaluation of 5- & 10-sample pooling. Indian J Med Res 2020; 152(1 & 2): 88–94. DOI: 10.4103/ijmr.IJMR_2304_20.

11. Ben-Ami R, Klochendler A, Seidel M, et al. Large-scale implementation of pooled RNA extraction and RT-PCR for SARS-CoV-2 detection. Clinical Microbiology and Infection 2020; 26(9): 1248–53. DOI: 10.1016/j.cmi.2020.06.009.

12. Cherif A, Grobe N, Wang X, Kotanko P. Simulation of Pool Testing to Identify Patients With Coronavirus Disease 2019 Under Conditions of Limited Test Availability. JAMA Network Open 2020; 3(6): e2013075–e. DOI: 10.1001/jamanetworkopen.2020.13075.

13. Bukhari SUK, Khalid SS, Syed A, Shah SSH. Smart Pooled sample Testing for COVID-19: A Possible Solution for Sparsity of Test Kits. medRxiv 2020: 2020.04.21.20044594. DOI: 10.1101/2020.04.21.20044594.

14. Žilinskas J, Lančinskas A, Guarracino MR. Pooled testing with replication: a mass testing strategy for the COVID-19 pandemics. medRxiv 2020: 2020.04.27.20076422. DOI: 10.1101/2020.04.27.20076422.

15. Regen F, Eren N, Heuser I, Hellmann-Regen J. A simple approach to optimum pool size for pooled SARS-CoV-2 testing. International Journal of Infectious Diseases 2020; 100: 324–6. DOI: 10.1016/j.ijid.2020.08.063.

16. Mutesa L, Ndishimye P, Butera Y, et al. A pooled testing strategy for identifying SARS-CoV-2 at low prevalence. Nature 2020. DOI: 10.1038/s41586-020-2885-5.

17. Hanel R, Thurner S. Boosting test-efficiency by pooled testing for SARS-CoV-2—Formula for optimal pool size. PLOS ONE 2020; 15(11): e0240652. DOI: 10.1371/journal.pone.0240652.

18. Aprahamian H, Bish DR, Bish EK. Optimal risk-based group testing. Management Science 2019;65(9):4365–84. DOI: 10.1287/mnsc.2018.3138.

19. Aprahamian H, Bish DR, Bish EK. Optimal Group Testing: Structural Properties and Robust Solutions, with Application to Public Health Screening. INFORMS Journal on Computing 2020. DOI: 10.1287/ijoc.2019.0942.

20. Corless RM, Gonnet GH, Hare DEG, Jeffrey DJ, Knuth DE. On the LambertW function. Advances in Computational Mathematics 1996; 5(1): 329–59. DOI: 10.1007/BF02124750.

21. Woloshin S, Patel N, Kesselheim AS. False Negative Tests for SARS-CoV-2 Infection — Challenges and Implications. New England Journal of Medicine 2020. DOI: 10.1056/NEJMp2015897.

22. Singh AK, Nema RK, Joshi A, et al. Evaluation of pooled sample analysis strategy in expediting case detection in areas with emerging outbreaks of COVID-19: A pilot study. PLOS ONE 2020; 15(9): e0239492. DOI: 10.1371/journal.pone.0239492.

23. Larremore DB, Wilder B, Lester E, et al. Test sensitivity is secondary to frequency and turnaround time for COVID-19 surveillance. medRxiv 2020: 2020.06.22.20136309. DOI: 10.1101/2020.06.22.20136309.

24. Woloshin S, Patel N, Kesselheim AS. False Negative Tests for SARS-CoV-2 Infection — Challenges and Implications. New England Journal of Medicine 2020; 383(6): e38. DOI: 10.1056/NEJMp2015897.

25. Shook N. Eight positive cases in latest COVID-19 testing data from NFL, NFLPA. September 13, 2020 2020. https://www.nfl.com/news/eight-positive-cases-in-latest-covid-19-testing-data-from-nfl-nflpa2020 (accessed October 19, 2020).

26. UIUC. Past seven-day case positivity rate. https://go.illinois.edu/COVIDTestingData (accessed November 16, 2020).

27. World Health Organization. Report of the WHO-China Joint Mission on Coronavirus Disease 2019 (COVID-19), 2020. https://www.who.int/docs/default-source/coronaviruse/who-china-joint-mission-on-covid-19-final-report.pdf.

28. Böger B, Fachi MM, Vilhena RO, Cobre AF, Tonin FS, Pontarolo R. Systematic review with meta-analysis of the accuracy of diagnostic tests for COVID-19. American Journal of Infection Control 2020. DOI: 10.1016/j.ajic.2020.07.011.

## References

1. Aprahamian H, Bish DR, Bish EK. Optimal Group Testing: Structural Properties and Robust Solutions, with Application to Public Health Screening. INFORMS Journal on Computing 2020; DOI: 10.1287/ijoc.2019.0942.

